# Effects of Age on the Association between Down Syndrome and Common Disease Conditions

**DOI:** 10.64898/2026.09.02.26362053

**Authors:** Akaninyene Noah, Hannah F Tavalire, Veronica Fitzpatrick, Hannah Graham, Shelly Verma, Laura Krohn, Katie Frank, Daniel Vockeroth, Brian Chicoine

## Abstract

**Importance:** Age-based screening and prevention guidelines in healthcare are developed largely in populations without Down syndrome (DS). If disease risk among individuals with DS differs significantly relative to typically developing (TD) individuals of similar ages, standard guidelines may overlook critical windows of care.

**Objective:** To determine the age-specific risk of ten common conditions in individuals with DS compared to a matched TD cohort to evaluate the presence of risk modification by age.

**Design:** Retrospective longitudinal cohort study.

**Setting:** Midwest healthcare system.

**Participants:** 4,912 individuals with DS and 37,924 TD controls, with clinical encounters between January 2005 and May 2025. Total person years (PYs) were 25,896 and 149,078 respectively.

**Exposure:** DS status

**Main Outcomes and Measures:** Incidence of acute sinusitis, anxiety, asthma, depression, gastroesophageal-reflux disorder, nervous system disorders, respiratory failure, thyroid gland disorders, vasomotor/allergic rhinitis, and vitamin-D deficiency. Incidence rate ratios (IRR) trajectories were analyzed using generalized additive mixed models. Wald tests assessed the presence of age modification.

**Results:** In our DS cohort (median_age_: 24, 48% female), we found evidence of age-related risk modification in 6 of 10 conditions examined. The relative incidence of anxiety was significantly lower among individuals with DS in early childhood (at age 5, IRR 0.23, 95%CI 0.19-0.29), compared to TD controls of the same age. However, risk in the DS cohort increased with age approaching equal levels with controls around ages 65. Similar trends were observed for depression. The association between DS and gastroesophageal disease was largely null except between ages 1–8, where the incidence among DS cohort was elevated (5–7 cases per 1000 PYs) compared to controls (2–3 cases per 1000 PYs). The association between DS and respiratory failure was statistically insignificant until age 53-70, where incidence among individuals with DS rose sharply (13-30 cases per 1000 PYs), while for TD counterparts it remained constant at 8-9 cases per 1000 PYs. Lastly, risk of vitamin-D deficiency fluctuated with a peak at age 20 (IRR 2.34, 95%CI2.02-2.72).

**Conclusions and Relevance:** These findings demonstrate age effect modification among individuals with DS across a variety of disease conditions and highlight the importance of population-specific care guidelines.

**Key Points:** *Question:* Does risk of disease vary significantly between individuals with Down syndrome and typically developing individuals by age?

*Findings:* Risks of anxiety, depression, thyroid gland disease, gastroesophageal disease, respiratory failure, and vitamins D deficiency varied significantly by age among individuals with Down syndrome, compared to a typically developing matched cohort.

*Meaning:* This manuscript describes key differences in age trajectories for different conditions experienced by individuals with Down syndrome. Previous disconnects in our understanding of age at diagnosis have likely led to mistimed and/or missed screening and prevention opportunities in individuals with Down Syndrome when following guidelines for typically developing counterparts.

## Introduction

Down syndrome (DS) occurs in approximately 1 in 700 live births in the US^1–3^ and is the most common chromosomal condition nationwide.^2^ Life expectancy for individuals with DS has risen from 25 years in 1983 to approximately 60 years in the 2020s.^4^ Individuals with DS experience accelerated biological aging relative to their typically developing counterparts.^5^ Prior studies have identified causal pathways linking the dysregulation of genes on the 21^st^ chromosome to increased cellular DNA damage,^6^ accelerated cognitive decline,^7^ and autoimmune hyperactivity.^8^ However, the overexpressed genes on chromosome 21 may offer protective effects against certain diseases, including various solid tumors^9^ and certain cardiovascular diseases, including hypertension.^10^ Several studies have observed interactions between age and genetic mutations in typically developing individuals that modify disease risk.^11,12^ Notable examples include *apolipoprotein E 4 (APOE)* and Alzheimer disease,^11^ *BRCA1/2* and breast cancer,^13^ and *Chromosome 9p21.3* and coronary artery disease.^14^ Likewise, studies have found sex-based interactions with genetic mutations on the risk of major depressive disorder,^12^ Alzheimer disease,^15^ and glioma.^12^ In individuals with DS, age-specific disease patterns have been described for dementia^16^. However, characterizations of age-related risk patterns for other common disease conditions affecting individuals with DS are limited.

With the rapidly growing population of adults with DS,^3^ developing tailored care guidelines for this population is critical. Several screening and care recommendation guidelines for clinicians are age-based and were largely developed for typically developing individuals. Whether this default set of guidelines misses critical care windows among those with DS is unclear. To address this gap, we investigated whether age influences the risk of various conditions in individuals with DS compared to a similar cohort without DS. Additionally, we aimed to identify differences in risk trajectories for a range of common disease conditions affecting individuals with DS to clarify critical windows for care interventions.

## Methods

### Study Setting

We conducted a retrospective cohort study using healthcare encounter data from a large healthcare system in the Midwest. The system consists of 25 hospitals, over 500 care locations, and the largest specialty care center for adults with DS in the US. The healthcare system’s Institutional Review Board approved the study protocol (IRB #00132715) and waived the informed consent requirement due to the retrospective nature of the study. This study followed the Strengthening the Reporting of Observational Studies in Epidemiology (STROBE) guidelines for observational studies.

### Study Population and Exclusion Criteria

Individuals with DS were identified using an International Classification of Diseases, 9^th^ revision, Clinical Modification (ICD-9-CM) code of 758.0 or an equivalent ICD-10-CM code (Q90). Between January 2005 and May 2025, we identified 5,926 individuals with DS. Controls were matched to the DS cohort at a 10:1 ratio based on age at first encounter, sex, race/ethnicity, year of first encounter, and encounter frequency during year of first encounter, yielding 59,832 controls.

We applied a set of exclusion criteria (**Figure 1**) to minimize potential sources of bias in our study. To ensure we did not include prevalent cases in our incidence estimates, we established a 1-year washout period such that patients were assigned an index date of 365 days after their first encounter. Those with less than 1 year of observation time or who had no encounters after their index date were excluded. After applying all exclusions, we had a baseline study population of 42,826 patients at index visit. For each diagnostic condition, if a patient had been diagnosed during the 1-year washout period, they were considered a prevalent case and removed from the at-risk analyses’ population for that condition. The at-risk analyses populations for each condition ranged from 40,487 to 42,098.

**Figure 1.**
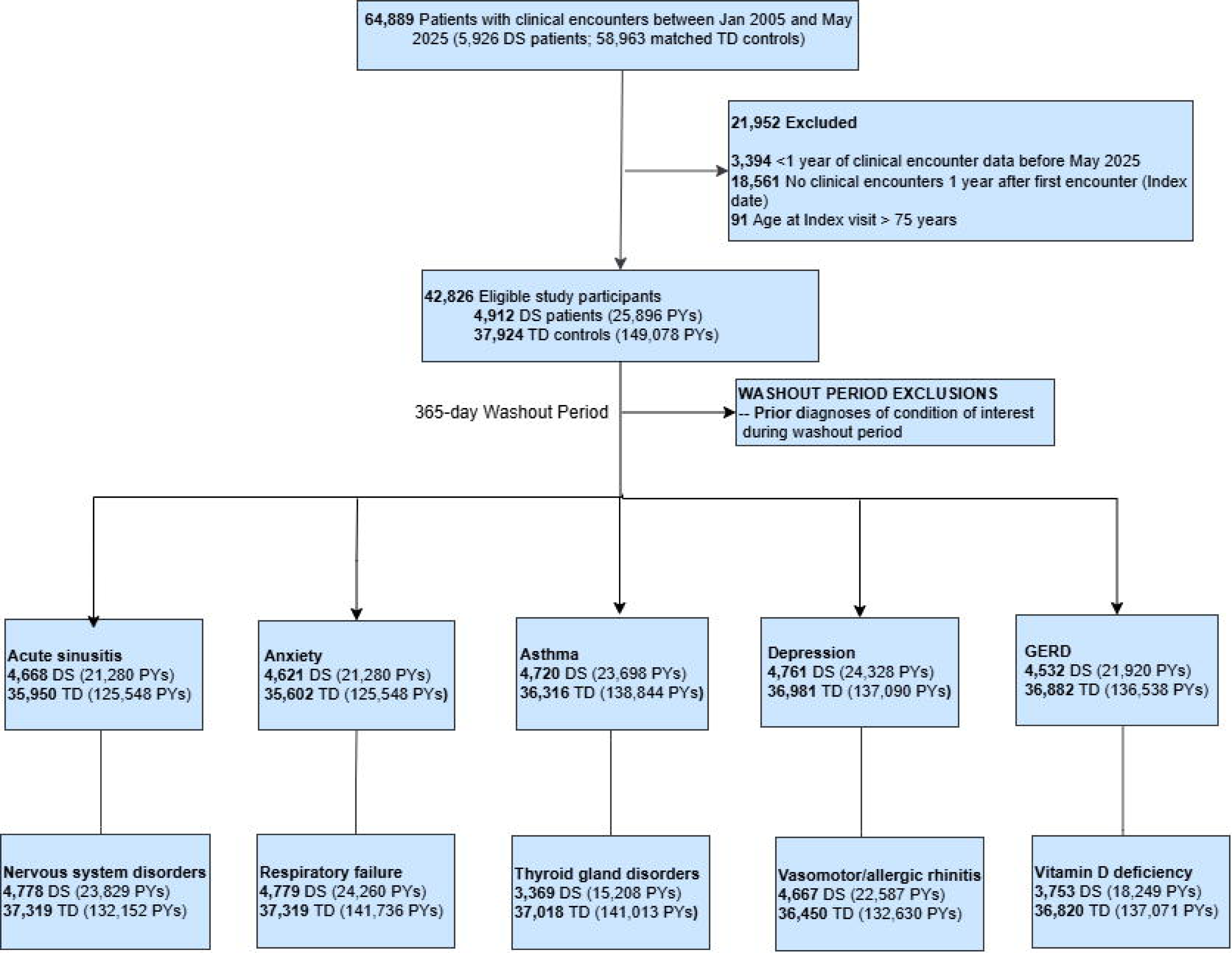
Inclusion/exclusion criteria for study population. Each diagnosis condition was analyzed individually, this the at-risk populations varied across conditions.

### Outcomes

A diagnosis condition was considered present if a patient had an encounter with the relevant ICD-10-CM code. We identified the ten most prevalent conditions among our baseline DS study population that also had a 5% prevalence within our baseline control group. Selected diagnoses conditions included: Acute sinusitis (J01), Anxiety disorder (F41), Asthma (J45), Depressive episodes (F32), Thyroid gland disorders (E00-E07), Gastroesophageal reflux disease (K21), Nervous system disorders (G89-G99), Respiratory failure (J96-J99), Vasomotor and allergic rhinitis (J30), and Vitamin D deficiency (E55). For each condition, at risk patients were followed until the date of their first incident diagnoses, the date of their last recorded encounters, or the study end date, whichever occurred first.

### Covariates

We collected both demographic and time-varying covariates measured at the end of each person-year of follow-up. Only person years in which a patient had at least one encounter were included. Fixed demographic variables included race/ethnicity (non-Hispanic [NH] White, NH Black, NH Asian, NH other race/ethnicity, unknown race/ethnicity), sex (male/female), and insurance type at index visit (commercial/private, Medicaid, Medicare, self-pay/other). Time-varying variables included age at the end of the calendar year, calendar year, and log-transformed total encounters within the calendar year as a proxy for surveillance intensity.

### Statistical Analyses

Baseline continuous variables were summarized using medians and interquartile ranges (IQRs) and compared using Wilcoxon rank sum tests. Categorical variables are presented using frequencies and percentages, with comparisons made using the Chi-square test. A two-sided p-value of less than 0.05 was considered statistically significant. Incidence was estimated as the number of new diagnoses per 1000 PYs at risk.

We utilized a Poisson generalized additive mixed model^17^ (GAMM) to estimate adjusted incidence rates (IR), incidence rate ratios (IRR), and absolute rate differences (RD). This model was selected because it can describe nonlinear trends over time while accounting for repeated observations among patients. We utilized a thin-plate spline^18^ for each continuous variable included in the model to capture the intrinsic nature of the association rather than pre-specifying an explicit form (i.e., allowing for nonlinear associations). Our fully adjusted model included DS status, age, a by-variable smooth function for DS status and age, sex, race/ethnicity, insurance type at index visit, calendar year, and log total encounters within the calendar year. A random intercept for each patient was also included. Smooth terms were estimated via restricted maximum likelihood (REML)^19^, and each model was tested using the *gam.check*^20^ function from the *mgcv* package in R to verify the adequacy of the k-basis dimensions of the smooth terms.^21^ All p-values for the k-indexes of each smooth term were above 0.05, indicating no evidence of overfitting. A Wald test^19^ was used to assess the significance of the by-variable smooth function of DS status and age, to quantify whether the difference between the DS and control group smooths were statistically different.^22^ False discovery rate (FDR) correction was used to account for multiple testing. We estimated predicted IR, IRR, and RDs using the 1^st^ to 99^th^ percentile of age values in the at-risk population to ensure we had sufficient sample size at each age.

Furthermore, we employed g-computation^23^ to estimate the marginal IRR of the association between DS and each diagnosis across all ages.^24,25^ This approach allowed us to evaluate an average estimate of the effect of DS on each diagnosed condition while also tracking how each estimate changes with age, using the same Poisson GAMM. Lastly, as a sensitivity analysis we expanded the washout period of each diagnosed condition to 2 years to exclude any prevalent cases the 1-year period may have missed. An association was considered significant if the 95% confidence interval for a given estimate did not include 1. All analyses were conducted using R software, version 4.5.^26^

## Results

### Demographics Characteristics

The baseline study population consisted of 4,912 individuals with DS and 37,924 controls, and together they contributed 25,896 and 149,078 person-years of follow-up, respectively (**Table 1**). Both groups had similar distributions of age at index date, sex, and race/ethnicity; however, there was a significant difference in the type of insurance at the index visits, with the DS cohort utilizing Medicare and Medicaid at higher rates, and the control group using self-pay at a higher rate. Individuals with DS per person contributed slightly more person-years (5 vs 3, respectively).

**Table 1.** Baseline characteristics of study population by DS status.

| Table 1. Baseline characteristics of study population by DS status |  |  |  |
| --- | --- | --- | --- |
|  | Individuals with DS | TD Controls | *P value* |
| No. of participants | 4,912 | 37,924 |  |
| Age, median (IQR) | 24 (11, 39) | 24 (11, 41) | <.001 |
| Follow up in years, median, (IQR) | 5 (3, 6) | 3 (2, 5) | <.001 |
| Total person years | 25,896 | 149,078 |  |
| Age groups across person years, No. (%) |  |  |  |
| 0 – 4 | 1,659 (6.4) | 4,596 (3.1) | <.001 |
| 5 – 17 | 5,189 (20.0) | 38,797 (26.0) |  |
| 18 – 24 | 3,505 (13.5) | 17,374 (11.65) |  |
| 25 – 49 | 11,284 (43.6) | 56,963 (38.2) |  |
| 50 – 64 | 3,876 (15.0) | 26,856 (18.0) |  |
| 65 + | 383 (1.5) | 4,472 (3.0) |  |
| Female sex, No. (%) | 2,389 (48.6) | 18,735 (49.4) | 0.3 |
| Race/ethnicity, No. (%) |  |  |  |
| Hispanic | 742 (15.1) | 5,704 (15.0) | 0.057 |
| NH Black | 448 (9.1) | 3,325 (8.8) |  |
| NH Other | 206 (4.2) | 1,497 (4.0) |  |
| NH White | 3247 (66.1) | 24,915 (65.7) |  |
| Unknown | 269 (5.5) | 2483 (6.6) |  |
| Insurance type at index visit, No. (%) |  |  |  |
| Commercial/Private | 1878 (38.1) | 26,221 (69.1) | <.001 |
| Medicaid | 1,258 (25.6) | 6,098 (16.1) |  |
| Medicare | 1,498 (30.5) | 914 (2.4) |  |
| Self-pay/Other | 284 (5.8) | 4,691 (12.4) |  |
| Abbreviation: DS, Down syndrome; IQR, Interquartile range |  |  |  |

### Age Effect Modification

We observed evidence of a significant interaction between DS status and age for six of the ten diagnosed conditions analyzed: anxiety disorder (P_fdr_<0.001), depressive episodes (P_fdr_ = 0.003), gastroesophageal reflux disease (P_fdr_ = 0.035), thyroid gland disorders (P_fdr_ < 0.001), respiratory failure (P_fdr_ =0.001), and vitamin D deficiency (P_fdr_ = 0.002) (**Figure 2; eFigure 1**)

**Figure 2.**
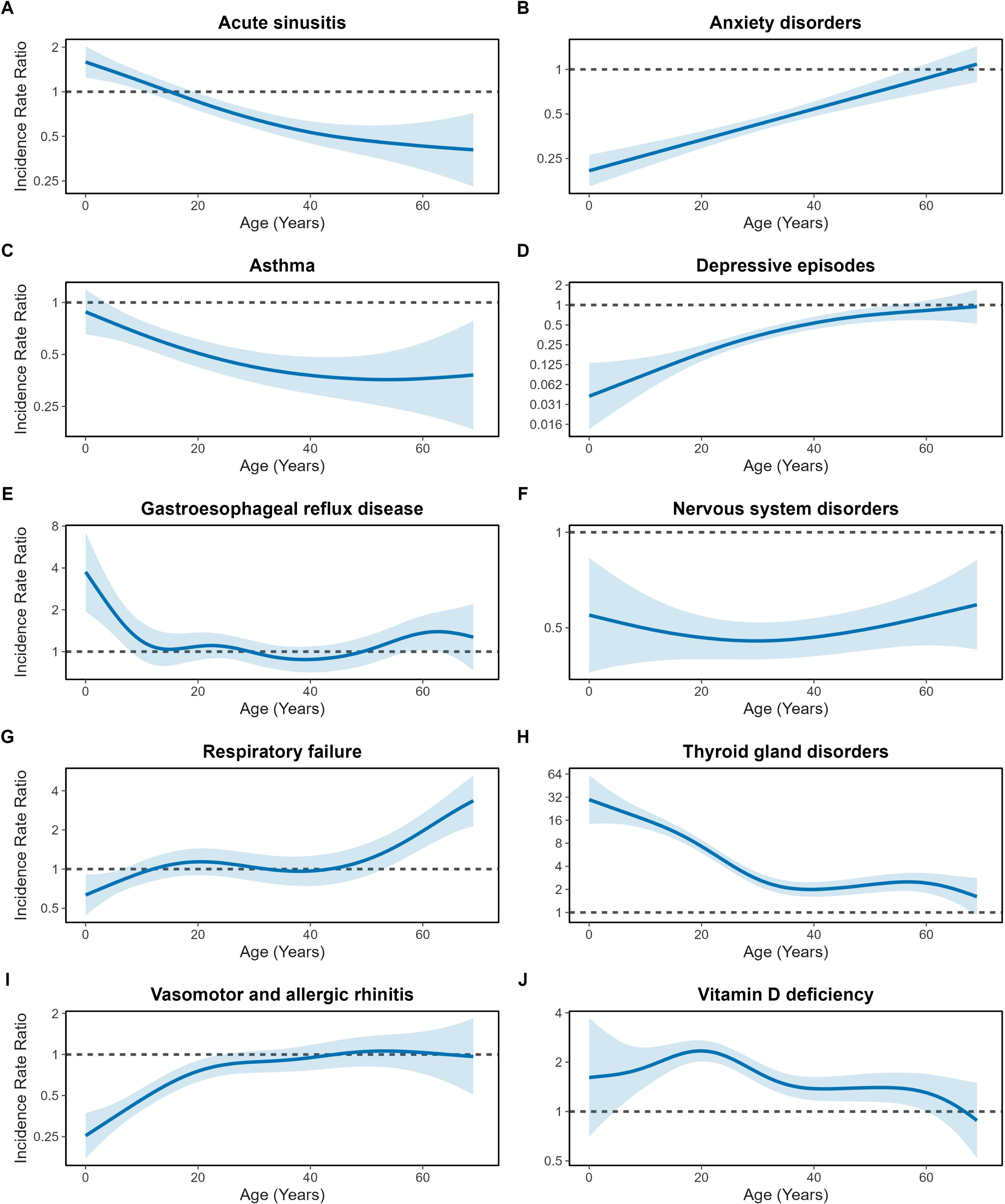
Age varying incidence rate ratio [IRR] for each diagnosed condition (A-J). The blue line represents the predicted IRR, and shaded area represents the 95% confidence interval. FDR-corrected Wald test p-value of the by-smooth DS status and age: A) Acute Sinusitis, *P_fd_*_r_ = 0.867; B) Anxiety disorder, *P_fd_*_r_<.001; C) Asthma, *P_fd_*_r_ = 0.215; D) Depressive episodes, *P_fd_*_r_ = 0.003; E) Gastroesophageal reflux disease, *P_fd_*_r_ = 0.035; F) Nervous system disorders, *P_fd_*_r_ = 0.616, G) Respiratory failure, *P_fd_*_r_ = 0.001; H) Thyroid gland disorders, *P_fd_*_r_ <.001; I) Vasomotor and allergic rhinitis, *P_fd_*_r_ = 0.084 J) Vitamin D deficiency, *P_fd_*_r_ = 0.002

### Anxiety disorder

The risk of being diagnosed with an anxiety disorder in individuals with DS is significantly influenced by age (**Table 2**; **Figure 2-3**). In early childhood (ages 1-8), the incidence of anxiety disorders is notably lower in individuals with DS compared to typically developing control of the same rate. The rate difference curve reaches its lowest point around age 21, showing approximately 43 fewer cases of anxiety disorders per 1000 PYs among individuals with DS (**Figure 3**). This trend appears to persist throughout adolescence and into late adulthood; however, the risk steadily increases with age, until the IRR reaches 1 around age 66 (IRR 1.01, 95% CI 0.78-1.31).

**Figure 3.**
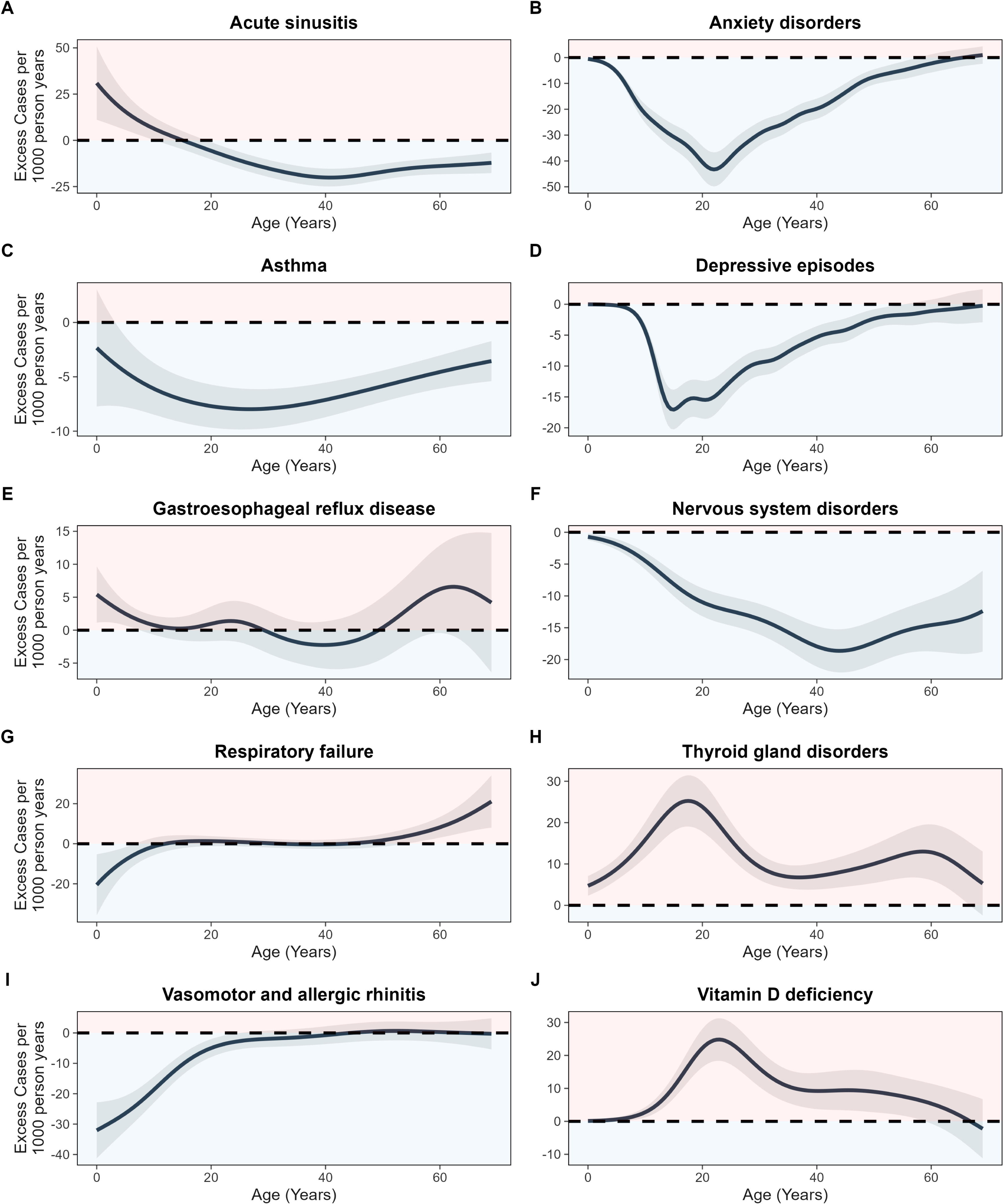
Age varying rate difference (RD) for each selected diagnoses condition (A-J). Red zones represent ages with excess relative cases (Increased risk) among individuals with DS and blue zones represent ages with fewer relative cases (decreased risk) among individuals with DS, compared to typically developing controls.

**Table 2.**
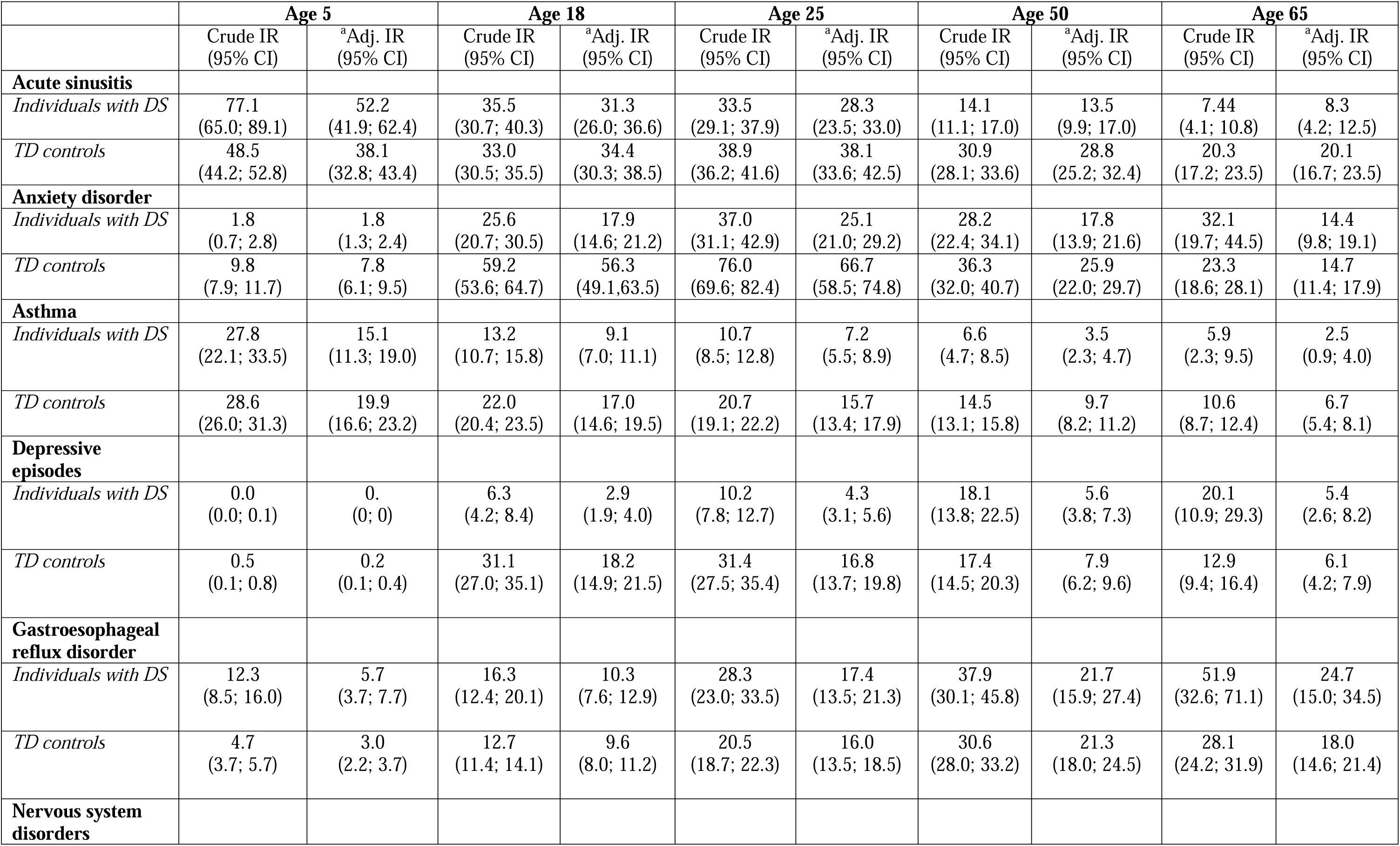
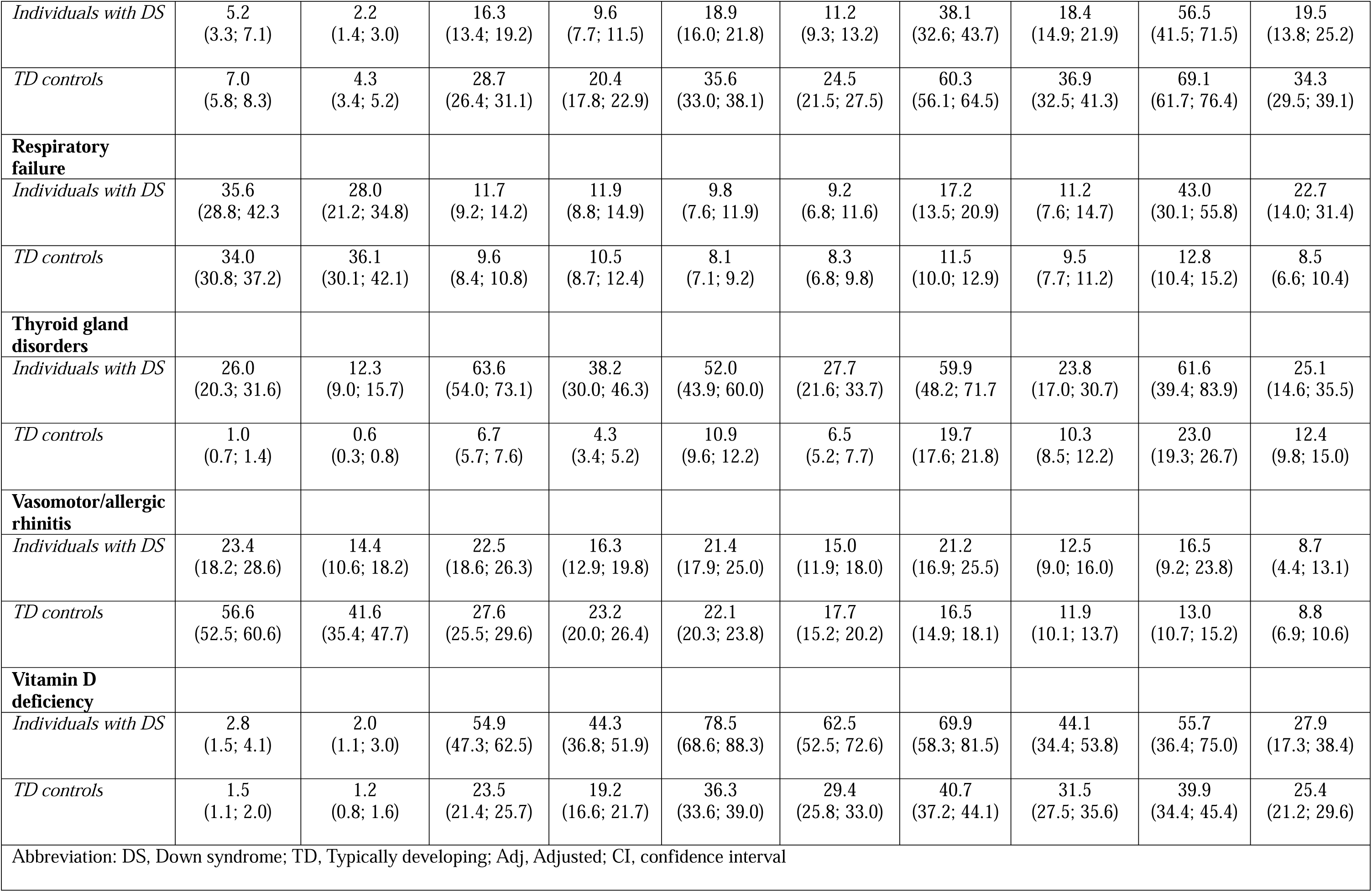
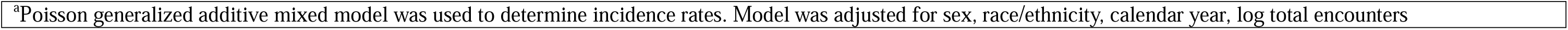
Incidence Rate per 1000 Person-Years, by Down Syndrome Status at Key Ages.

### Depressive episodes

Similar to anxiety disorders, the risk of being diagnosed with a depressive episode was substantially lower among individuals with DS in early childhood (0 – 0.1 cases per 1000 PYs), compared to typically developing individuals of the same age (0.2 – 1.4 cases per 1000 PYs). The low point of the rate difference curve was around age 15, with about 15 fewer cases among individuals with DS per 1000 PYs. We also observed the same trend, where risk of depressive episodes among the DS cohort appeared to increase with increasing age, with the confidence interval crossing 1 around age

### Gastroesophageal reflux disease

The 95% confidence interval of marginal IRR for the association between DS status and gastroesophageal reflux disease crosses 1, which would suggest a null or statistically insignificant association with DS status (IRR_m_ 1.08, 95% CI 0.95-1.12). However, this masks a critical window between risk ages 1 – 8, where the risk is significantly elevated among individuals with DS (at age 1: IRR 3.27, 95%CI 1.82-5.88; age 8: IRR 1.39, 95% CI 1.01-1.91) (**Figure 2**). After this age window, the association with DS remains at or close to 1 throughout the lifespan.

### Respiratory failure

Here the marginal IRR (1.11, 95% CI 0.97-1.28) masks two key age windows among individuals with DS (**eFigure 1**). In early childhood, between ages 1-6, individuals with DS had a reduced risk of respiratory failure relative to typically developing controls of the same age (at age 3: IRR 0.71, 95% CI 0.55-0.92). The association remained statistically insignificant until age 53, where the incidence among individuals with DS rose sharply until age 70 (13-30 cases per 1000 PYs), while remaining constant for controls within the same age range (8-9 cases per 1000 PYs) **(Figure 2; eFigure 2**)

### Thyroid gland disorders

Individuals with DS have a substantial higher risk of thyroid gland disorders in early childhood (at age 5: IRR 22.0, 95% CI 14.3-33.6). This pattern continuous throughout the lifespan, from adolescence to late adulthood, but declines with age (**Figure 2**). The rate difference curve shows bimodal pattern with peaks around age 18 (excess of 25 cases per 1000 PYs, 95% CI 19-31), and another around age 58 (excess cases per 1000 PYs: 13, 95% CI 6-20) (**Figure 3**)

### Vitamin D disorder

The association between DS status and vitamin D disorder appeared to fluctuate widely across age with a peak around age 21 corresponding to an elevated IRR of 2.33 (95% CI: [1.99, 2.70]) and an excess case count among the DS cohort of 24 cases per 1000 PYs (**Figure 3**)

### Sensitivity analyses

Using a 2-year washout period reduced our baseline study population at index to 36,726 (4,436 individuals with DS; 32,289 controls). Our at-risk populations across all diagnosed conditions ranged from 33,584 to 35,785. We still observe evidence of effect modification by age in 5 out of 10 conditions; anxiety disorder, depressive episodes, respiratory failure, thyroid gland disorders, and vitamin D disorder. (**eFigure 3**)

## Discussion

Our findings suggest complex interactions between DS status and age across multiple conditions. The incidence of thyroid gland disorders was significantly elevated among individuals with DS, largely in early years, and decreased but remained elevated with age. Whereas the risks of both anxiety and depressive episodes were substantially lower among individuals with DS in childhood and rose steadily with age. The excess risk associated with gastroesophageal reflux disease was confined to ages 1 to 8. Respiratory failure showed no meaningful difference between both groups until age 53, at which point the incidence among individuals with DS rose sharply, while among typically developing individuals it remained constant. The excess risk associated with gastroesophageal reflux disease was confined to ages 1 to 8. Respiratory failure showed no meaningful difference between both groups until age 53, at which point the incidence among individuals with DS rose sharply, while among typically developing individuals it remained constant. Finally, the incidence of vitamin D deficiency varied with age, peaking in early childhood in individuals with DS.

The risk patterns identified here broadly align with existing DS literature.^27–29^ The late spike in respiratory failure incidence beginning around the median life expectancy for individuals with DS aligns with morbidity trends seen in individuals with DS.^29^ Respiratory illnesses, including aspiration pneumonia and pneumonitis, are leading causes of mortality among adults with DS.^29^ Additionally, impaired respiratory functioning is associated with an increased risk of Alzheimer’s dementia.^30^ A spike in respiratory illness around 53-55 aligns with the average age of onset of Alzheimer’s disease in the DS population.

Gastroesophageal reflex disorder is a common issue for children with DS.^31^ This condition has been linked to reduced sphincter tone between the esophagus and stomach.^31–33^ Dietary choices, such as the consumption of caffeinated beverages like soda, may exacerbate this issue. Additionally, developmental delays in achieving an upright posture while eating could contribute to the higher incidence in this age group. Low vitamin D levels have been linked to increased autoimmune disorders,^34^ which are intrinsically associated with overexpressed genes on chromosome 21^8^. Studies among children with DS have noted lower counts of CD3, CD56 and CD16 cells^35^, and granulocytes^36^, which could lead to our observed higher incidence of vitamin D deficiency in early childhood.

The converging trajectories observed for depressive episodes and anxiety disorders present two interpretations. The incidence of these conditions may in fact be lower in childhood and adolescence and rise with age among the DS population. Alternatively, psychiatric conditions in younger individuals with DS may be attributed to the syndrome itself rather than recorded as a distinct diagnosis. Symptoms such as withdrawal or changes in behavior may be attributed to other issues due to diagnostic overshadowing resulting in underestimation by diagnostic codes for individuals with DS. ^37,38^ This would impact younger DS patients more, as communication delays could hinder the identification of mental health problems.

### Implications for Clinical Practice

Overwhelmingly, screening and care recommendation guidelines for clinicians are age-based and were largely developed using clinical observations of typically developing individuals. This study provides evidence that critical screening and prevention windows are likely to be missed or mistimed in individuals with DS when following existing care guidelines. Specifically, the extremely low relative incidences of generalized anxiety and depression among individuals with DS at younger ages suggest that the current tools might be underdiagnosing generalized anxiety and depression within the population.

The PHQ-2 and PHQ-9 are commonly used screening tools to detect depression and anxiety; however, these tools have not been validation for use in people with DS. For younger patients who experience communication difficulties, caregivers often complete these assessments, which may lead to underreporting of mental health issues. At the adult Down Syndrome clinic, a specialized DS center for adolescents and adults within the healthcare system our EHR data originated from, providers who specialized in DS rely on direct questioning of both caregivers and patients to diagnose depression and anxiety. Within our study population, we found that DS patients with at least one encounter with a DS specialized provider had a higher prevalence of anxiety diagnoses compared to those who did not meet with a DS specialized provider. (**eTable 1**).

### Limitations

This study has several limitations. First, the healthcare system from which we obtained data contains a large adult DS specialty care center, where early screenings for thyroid disorders and vitamin D deficiency are routine. The higher observed incidence may be reflected earlier, and more frequent testing rather than increased risk compared to the matched cohort. About 36% of our baseline DS cohort had at least one encounter at the DS specialty care center. We did not observe any significant difference in the prevalence of thyroid disorders by specialty care encounters, however patients with one or more specialty care encounters had a higher prevalence of vitamin D deficiency (54% vs. 12%) (**eTable 1**). Secondly, we used diagnosis codes as our key indicator for new-onset cases of a condition. Individuals who already have a condition and are referred to the healthcare system for treatment or further diagnostic tests would artificially inflate our count of incident cases. To mitigate this prevalent user bias, we varied our washout period between 1-2 years and observed the presence of effect modification by age within 5 of our 10 selected diagnosed conditions.

## Conclusions

These findings demonstrate age effect modification for risk of common conditions among individuals with DS and demonstrate the importance of population-specific care guidelines.

## Data Availability

All data produced in the present study are not available due to institutional restrictions of protected health information. Analytic code used to generate study findings are available upon request to authors.

## Author contributions

AN was responsible for study conceptualization and performed the statistical analysis. AN, VF, HFT, and DV contributed to the writing and editing of the original draft. BC supervised the project. HG, SV, and KF provided critical edits to the manuscript for important clinical content. All authors reviewed and approved the final draft.

## Conflicts of interest

The authors declare that they have no conflicts of interest to disclose.

## Acknowledgments

None

## Sources of funding and support

None

## Data Sharing Statement

Data are not publicly available due to institutional restrictions on protected health information. Analytic code used to generate study findings are available from corresponding author upon request.

## Notes

### Competing Interest Statement

The authors have declared no competing interest.

### Author Declarations

Advocate Health Institutional Review Board approved the study protocol (IRB #00132715) and waived the informed consent requirement due to the retrospective nature of the study.

## References

1. Mai CT, Isenburg JL, Canfield MA, et al. National population-based estimates for major birth defects, 2010–2014. Birth Defects Research. 2019;111(18):1420–1435. 10.1002/bdr2.1589

2. Centers for Disease Control and Prevention. Living with Down Syndrome. Updated November 22, 2024. Accessed January 26, 2026. https://www.cdc.gov/birth-defects/living-with-down-syndrome/index.html

3. de Graaf G, Buckley F, Skotko BG. Estimates of the live births, natural losses, and elective terminations with Down syndrome in the United States. American Journal of Medical Genetics Part A. 2015;167(4):756–767. 10.1002/ajmg.a.37001

4. Presson AP, Partyka G, Jensen KM, et al. Current Estimate of Down Syndrome Population Prevalence in the United States. The Journal of Pediatrics. 2013;163(4):1163–1168. doi:10.1016/j.jpeds.2013.06.013

5. Zigman WB. Atypical aging in Down syndrome. Dev Disabil Res Rev. 2013;18(1):51–67. doi:10.1002/ddrr.1128

6. Feaster WW, Kwok LW, Epstein CJ. Dosage effects for superoxide dismutase-1 in nucleated cells aneuploid for chromosome 21. Am J Hum Genet. Nov 1977;29(6):563–70.

7. Russo ML, Sousa AMM, Bhattacharyya A. Consequences of trisomy 21 for brain development in Down syndrome. Nat Rev Neurosci. Nov 2024;25(11):740–755. doi:10.1038/s41583-024-00866-2

8. Araya P, Waugh KA, Sullivan KD, et al. Trisomy 21 dysregulates T cell lineages toward an autoimmunity-prone state associated with interferon hyperactivity. Proc Natl Acad Sci U S A. Nov 26 2019;116(48):24231–24241. doi:10.1073/pnas.1908129116

9. Taherifard E, Taherifard E, Bouffet E, Satge D, Abdelbaki MS. Solid Tumor Incidence and Patterns in Individuals With Down Syndrome: A Systematic Review and Meta-Analysis. Pediatr Blood Cancer. Aug 2025;72(8):e31775. doi:10.1002/pbc.31775

10. Santoro JD, Lee S, Mlynash M, Mayne EW, Rafii MS, Skotko BG. Diminished Blood Pressure Profiles in Children With Down Syndrome. Hypertension. Mar 2020;75(3):819–825. doi:10.1161/HYPERTENSIONAHA.119.14416

11. Riedel BC, Thompson PM, Brinton RD. Age, APOE and sex: Triad of risk of Alzheimer’s disease. J Steroid Biochem Mol Biol. Jun 2016;160:134–47. doi:10.1016/j.jsbmb.2016.03.012

12. Ostrom QT, Kinnersley B, Wrensch MR, et al. Sex-specific glioma genome-wide association study identifies new risk locus at 3p21.31 in females, and finds sex-differences in risk at 8q24.21. Sci Rep. May 9 2018;8(1):7352. doi:10.1038/s41598-018-24580-z

13. Kuchenbaecker KB, Hopper JL, Barnes DR, et al. Risks of Breast, Ovarian, and Contralateral Breast Cancer for BRCA1 and BRCA2 Mutation Carriers. JAMA. 2017;317(23):2402–2416. doi:10.1001/jama.2017.7112

14. Ellis KL, Pilbrow AP, Frampton CM, et al. A common variant at chromosome 9P21.3 is associated with age of onset of coronary disease but not subsequent mortality. Circ Cardiovasc Genet. Jun 2010;3(3):286–93. doi:10.1161/CIRCGENETICS.109.917443

15. Farrer LA, Cupples LA, Haines JL, et al. Effects of age, sex, and ethnicity on the association between apolipoprotein E genotype and Alzheimer disease. A meta-analysis. APOE and Alzheimer Disease Meta Analysis Consortium. JAMA. Oct 22-29 1997;278(16):1349–56.

16. Boerwinkle AH, Gordon BA, Wisch J, et al. Comparison of amyloid burden in individuals with Down syndrome versus autosomal dominant Alzheimer’s disease: a cross-sectional study. Lancet Neurol. Jan 2023;22(1):55–65. doi:10.1016/s1474-4422(22)00408-2

17. Lin X, Zhang D. Inference in Generalized Additive Mixed Models by Using Smoothing Splines. Journal of the Royal Statistical Society Series B: Statistical Methodology. 1999;61(2):381–400. doi:10.1111/1467-9868.00183

18. Wood SN. Thin Plate Regression Splines. Journal of the Royal Statistical Society Series B: Statistical Methodology. 2003;65(1):95–114. doi:10.1111/1467-9868.00374

19. Wood SN. Fast Stable Restricted Maximum Likelihood and Marginal Likelihood Estimation of Semiparametric Generalized Linear Models. Journal of the Royal Statistical Society Series B: Statistical Methodology. 2011;73(1):3–36. doi:10.1111/j.1467-9868.2010.00749.x

20. Wood S. Generalized Additive Models: An Introduction With R. vol 66. 2006:391.

21. Wood SN. Stable and Efficient Multiple Smoothing Parameter Estimation for Generalized Additive Models. Journal of the American Statistical Association. 2004/09/01 2004;99(467):673–686. doi:10.1198/016214504000000980

22. Wood SN. On p-values for smooth components of an extended generalized additive model. Biometrika. 2013;100(1):221–228. doi:10.1093/biomet/ass048

23. Naimi AI, Cole SR, Kennedy EH. An introduction to g methods. Int J Epidemiol. Apr 1 2017;46(2):756–762. doi:10.1093/ije/dyw323

24. Almasi-Hashiani A, Nedjat S, Mansournia MA. Causal Methods for Observational Research: A Primer. Arch Iran Med. Apr 1 2018;21(4):164–169.

25. Wood SN. Inference and computation with generalized additive models and their extensions. TEST. 2020/06/01 2020;29(2):307–339. doi:10.1007/s11749-020-00711-5

26. R: A Language and Environment for Statistical Computing. R Foundation for Statistical Computing; 2025. https://www.r-project.org/

27. Santoro SL, Chicoine B, Jasien JM, et al. Pneumonia and respiratory infections in Down syndrome: A scoping review of the literature. American Journal of Medical Genetics Part A. 2021;185(1):286–299. 10.1002/ajmg.a.61924

28. Macchini F, Leva E, Torricelli M, Valadè A. Treating acid reflux disease in patients with Down syndrome: pharmacological and physiological approaches. Clinical and Experimental Gastroenterology. 2011/01/25 2011;4(null):19–22. doi:10.2147/CEG.S15872

29. Landes SD, Stevens JD, Turk MA. Cause of death in adults with Down syndrome in the United States. Disabil Health J. Oct 2020;13(4):100947. doi:10.1016/j.dhjo.2020.100947

30. Russ TC, Kivimäki M, Batty GD. Respiratory Disease and Lower Pulmonary Function as Risk Factors for Dementia: A Systematic Review With Meta-analysis. Chest. 2020/06/01/ 2020;157(6):1538–1558. 10.1016/j.chest.2019.12.012

31. Buchin PJ, Levy JS, Schullinger JN. Down’s syndrome and the gastrointestinal tract. J Clin Gastroenterol. 1986/04// 1986;8(2):111–114. doi:10.1097/00004836-198604000-00002

32. Mayo Clinic. Gastroesophageal reflux disease (GERD). May 20th 2026, https://www.mayoclinic.org/diseases-conditions/gerd/symptoms-causes/syc-20361940

33. Hillemeier C, Buchin PJ, Gryboski J. Esophageal Dysfunction in Down’s Syndrome. Journal of Pediatric Gastroenterology and Nutrition. 1982;1(1):101–104. 10.1002/j.1536-4801.1982.tb08302.x

34. Vincenzi F, Smirne C, Tonello S, Sainaghi PP. The Role of Vitamin D in Autoimmune Diseases. International Journal of Molecular Sciences. 2026;27(1):555.

35. de Hingh YC, van der Vossen PW, Gemen EF, et al. Intrinsic abnormalities of lymphocyte counts in children with down syndrome. J Pediatr. Dec 2005;147(6):744–7. doi:10.1016/j.jpeds.2005.07.022

36. Bloemers BL, van Bleek GM, Kimpen JL, Bont L. Distinct abnormalities in the innate immune system of children with Down syndrome. J Pediatr. May 2010;156(5):804–9, 809 e1-809 e5. doi:10.1016/j.jpeds.2009.12.006

37. Lazris A, Roth A, Haskell H, James J. Diagnostic Overshadowing: When Cognitive Biases Can Harm Patients. Am Fam Physician. Sep 2023;108(3):292–294.

38. Molloy R, Munro I, Pope N. Understanding the experience of diagnostic overshadowing associated with severe mental illness from the consumer and health professional perspective: a qualitative systematic review protocol. JBI Evidence Synthesis. 2020/11/07 2021;19(6):1362–1368. doi:10.11124/JBIES-20-00244

