## Supplemental Material for "Effects of Age on the Association between Down Syndrome and Common Disease Conditions"

| **eTable 1. Prevalence of Common Disease Condition among Individuals with DS, Stratified by DS Specialty Care** | | | | |
| --- | --- | --- | --- | --- |
|  |  | **Specialty care visits** | |  |
| **Condition** | **Overall DS at-risk population**  **n / N, (%)** | $\boldsymbol{\geq}$**1 DS Specialty care Visits**  **n / N (%)** | **0 DS Specialty Care Visits**  **n / N (%)** | ***P value*** |
| Acute sinusitis | 492 / 4668 (10.5) | 87 / 1808 (4.8) | 405 / 2860 (14.2) | <.001 |
| Anxiety disorder | 513 / 4621 (11.1) | 266 / 1637 (16.3) | 247 / 2984 (8.3) | <.001 |
| Asthma | 242 / 4720 (5.1) | 36 / 1795 (2.0) | 206 / 2925 (7.0) | <.001 |
| Depressive episodes | 228 / 4532 (4.8) | 73 / 1751 (4.2) | 155 / 3010 (5.1) | 0.13 |
| Gastroesophageal reflux disorder | 474 / 4532 (10.5) | 212 / 1668 (12.7) | 262 / 2864 (9.1) | <.001 |
| Nervous system disorders | 479 / 4778 (10.0) | 157 / 1777 (8.8) | 322 / 3001 (10.7) | 0.035 |
| Respiratory failure | 356 / 4779 (7.4) | 71 / 1846 (3.8) | 285 / 2933 (9.7) | <.001 |
| Thyroid gland disorders | 622 / 3369 (18.5) | 171 / 937 (18.2) | 451 / 2432 (18.5) | 0.800 |
| Vasomotor/allergic rhinitis | 407 / 4667 (8.7) | 138 / 1725 (8.0) | 269 / 2942 (9.1) | 0.200 |
| Vitamin D deficiency | 786 / 3753 (20.9) | 424 / 783 (54.2) | 362 / 2970 (12.2) | <.001 |
| P values obtained from chi-square test | | | | |

**eFigure 1.** Point estimates for marginal IRR

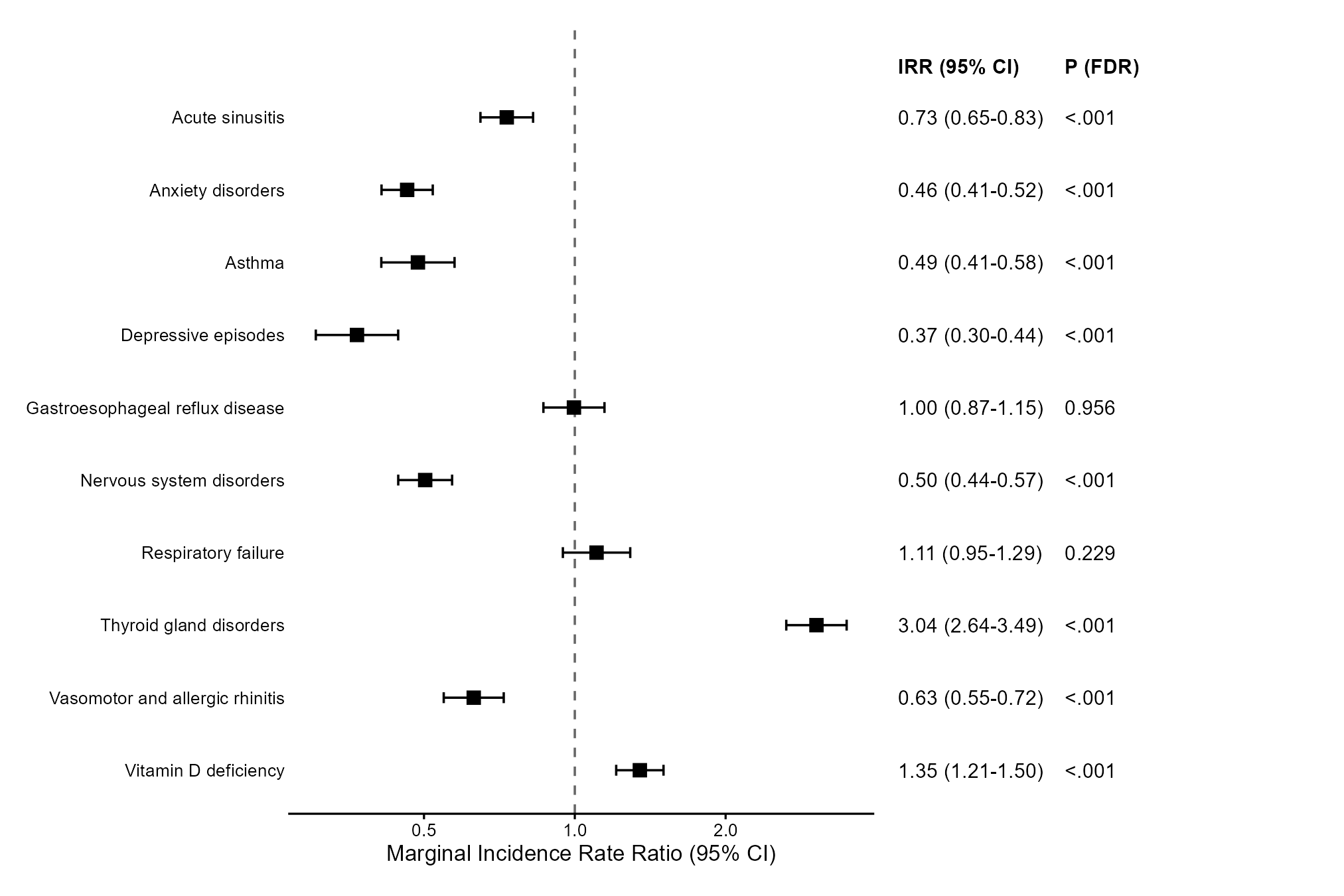

**eFigure 2.** Age varying incidence rates for each diagnosed condition (A-J). The blue line represents the incidence rate per 1000 person years for individuals with down syndrome, and orange line represents the incidence rate for typically developing controls.

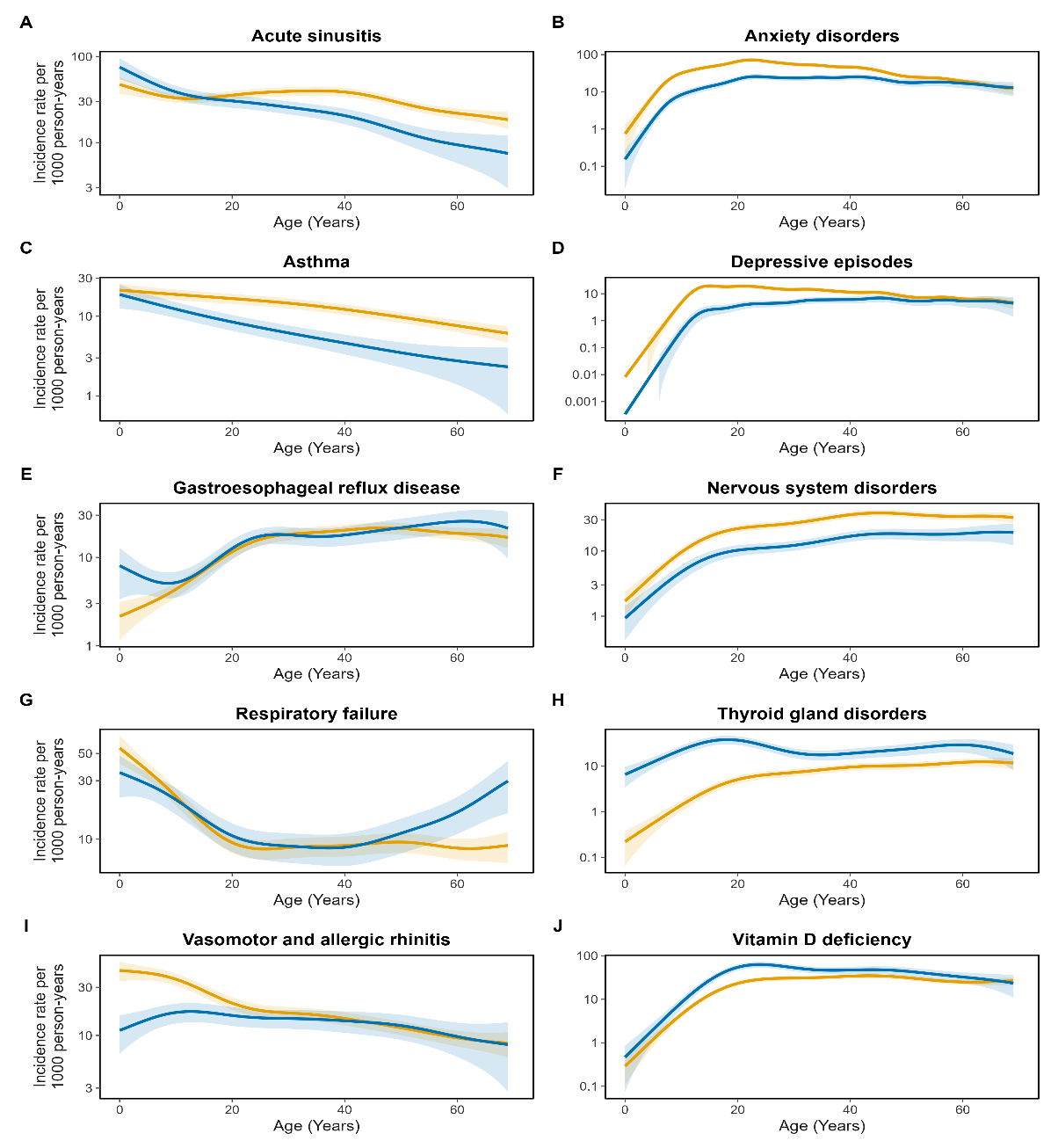

**eFigure 3.** Age varying incidence rate ratio using a 2-year washout period. The blue line represents the predicted IRR, and shaded area represents the 95% confidence interval.

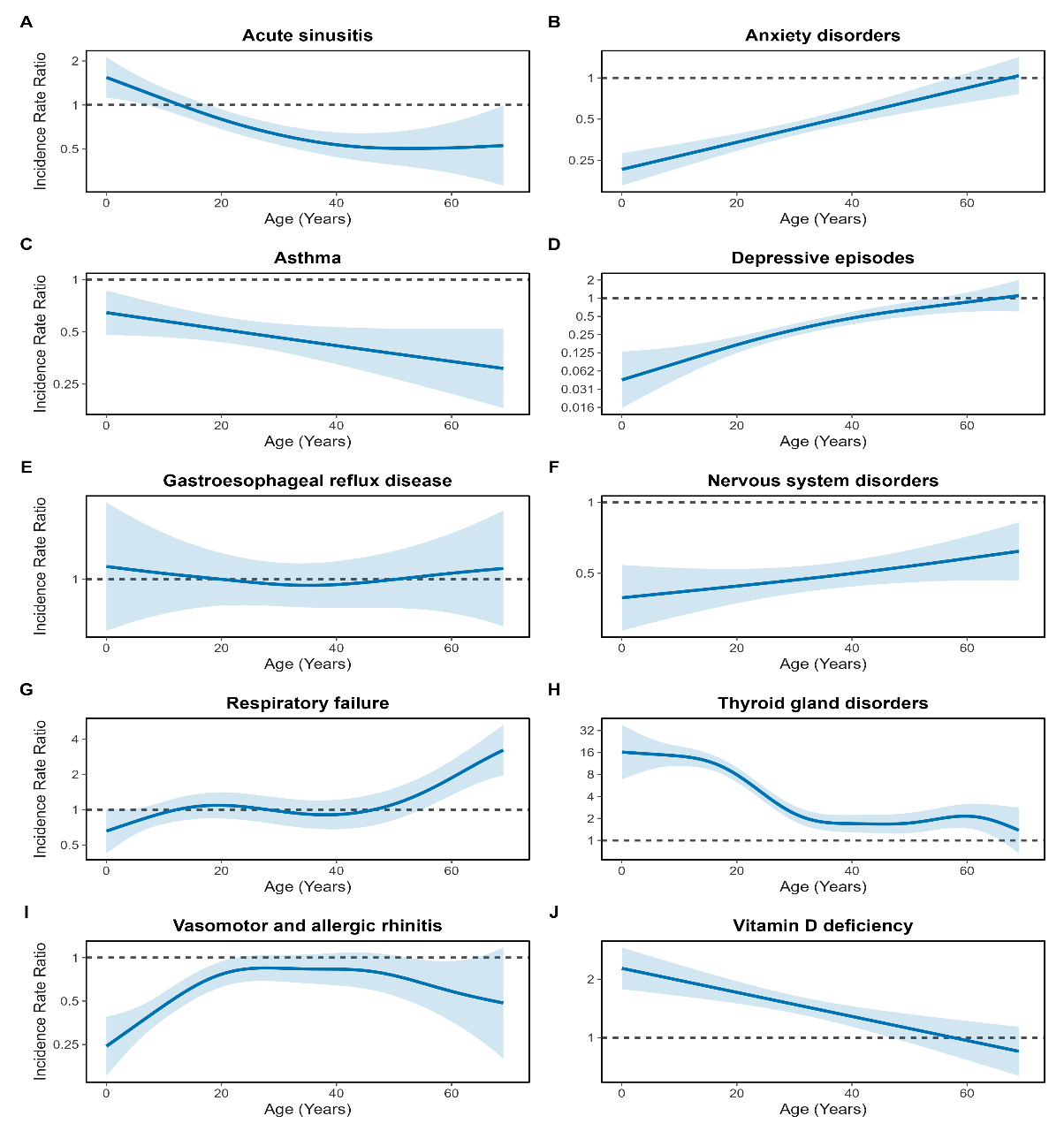
